# Geospatial Analysis of the Association between Medicaid Expansion, Minimum Wage Policies, and Alzheimer’s Disease Dementia Prevalence in the United States

**DOI:** 10.1101/2024.09.10.24313418

**Authors:** Abolfazl Mollalo, Sara Knox, Jessica Meng, Andreana Benitez, Leslie A. Lenert, Alexander V. Alekseyenko

## Abstract

**Background:** Previous studies indicate that improved healthcare access through Medicaid expansion and the alleviation of socioeconomic stressors via higher minimum wages lead to improved health outcomes. However, a significant gap remains in examining the distribution of Alzheimer’s disease (AD) dementia in the context of these policies. Thus, this study investigates the spatial relationships between the Medicaid expansion, minimum wage policy, and AD dementia prevalence across the US.

**Methods:** We employed the Getis-Ord Gi* statistic to identify hot spots and cold spots of AD dementia prevalence at the county level across the US. We compared these locations with the county-level overall social vulnerability index (SVI) scores. Moreover, we assessed the proportion of hot and cold spots at the state level based on Medicaid expansion and minimum wage status.

**Results:** Our analysis revealed that the most vulnerable SVI quartile (Q4) had the highest number of hot spots (n=311, 64.8%), while the least vulnerable quartile (Q1) had the fewest hot spots (n=22, 4.6%) (*χ*^2^=307.41, *P*<0.01). Additionally, Q4 had the fewest cold spots (n=54, 11.7%). States that adopted Medicaid expansion had a significantly lower proportion of hot spots compared to non-adopting states (*P*<0.05), and the non-adopting states had significantly higher odds of having hot spots than adopting states (OR=2.58, 95% CI: 2.04–3.26, *P*<0.001). Conversely, the non-adopting states had significantly lower odds of having cold spots compared to the adopting states (OR=0.24, 95% CI: 0.19–0.32, *P*<0.01). States with minimum wage levels at or below the federal level showed significantly higher odds of having hot spots than states with a minimum wage above the federal level (OR=1.94, 95% CI: 1.51–2.49, *P*<0.01).

**Discussion:** Our findings suggest significant disparities in AD dementia prevalence related to socioeconomic and policy factors and lay the groundwork for future causal analyses to examine whether these policies have impacted disparities in healthcare access for people with AD dementia across the US.

## 1. Introduction

Alzheimer’s disease (AD) dementia, an escalating public health concern, affected approximately 5.6 million US residents in 2019 [1]. With the aging of the baby boomer generation, this number is projected to increase significantly to 13.8 million by 2050 [2], highlighting the urgent need to improve healthcare access and address disparities. In response, various policies have been enacted to improve health coverage and care for underserved populations. One such policy in the US is Medicaid, a program jointly funded by federal and state governments, which provides healthcare coverage to individuals and families with limited income or resources [3]. The Affordable Care Act (ACA) expanded Medicaid eligibility to include all US residents with incomes up to 138% of the federal poverty line; however, each state can choose whether to adopt expansion [4]. As of July 14, 2023, 40 states and Washington DC have implemented this expansion [5]. Following this implementation, numerous studies investigated the impacts of Medicaid expansion on healthcare access and clinical outcomes. Findings consistently indicate that the expansion has led to lower uninsurance rates [6, 7], increased utilization of primary care and preventive services [8], decreased inpatient hospitalization rates [9], and reduced all-cause mortality [10].

Socioeconomic deprivation and neighborhood disadvantage are associated with an increased risk of dementia [11, 12]. Thus, it is essential to consider how US governmental policies that are linked to socioeconomics, such as the minimum wage, may influence the risk of dementia. Since 2009, the federal minimum wage has been set at $7.25 per hour. However, as of July 2024, 34 states have enacted higher minimum wages, with some, such as California, reaching up to $16 per hour [13].

Research on the impacts of minimum wage policies on health outcomes is limited and varied in results. For instance, increases in minimum wage have been associated with modest decreases in heart disease mortality [14] and improved healthcare access among White men, Black women, and Latina women [15]. Additionally, while minimum wage increases have been found to improve women’s health, they may adversely affect men’s health [16]. States with stagnant minimum wages experienced some of the poorest health outcomes, including higher rates of obesity and low birth rates [17].

Notably, prior research has shown that cumulative exposure to low wages in midlife is associated with faster memory decline in late life [18]. These findings underscore the critical need to consider socioeconomic factors in health policies. In this context, the Social Vulnerability Index (SVI) can be a valuable tool for assessing the level of disadvantage within neighborhoods [19].

Geospatial analysis has provided a robust framework for investigating the spatial relationship of Medicaid adoption or minimum wage policies with various health outcomes, such as cancer [20], sexually transmitted infections [21], and access to healthcare [22, 23]. Jemal et al. (2017) examined changes in uninsured rates and early-stage cancer diagnoses among nonelderly patients following the ACA implementation. Their results indicated a notable decrease in uninsured rates among newly diagnosed cancer patients across all income brackets in Medicaid expansion and non-expansion states. However, the most significant decrease was observed among low-income patients in expansion states compared to non-expansion states [24]. In a recent study, Li et al. (2024) utilized inpatient claims data to explore spatiotemporal patterns of hospitalizations involving comorbid cancer and dementia. They identified 22 hotspots and found that the likelihood of a county being a hotspot significantly increased with the percentage of dual Medicare Medicaid beneficiaries while decreasing with higher rurality [25]. The impact of minimum wage on health studies remains a relatively unexplored area of research [26] and is rarely explored from a geospatial perspective. Rath et al. (2022) examined the effects of implementing a minimum wage in Hong Kong on suicide rates and found that the introduction of the minimum wage led to an immediate 13.0% reduction in monthly suicide rates (*P*<0.05). Notably, there was a 15.8% immediate decline in suicide rates among older working-aged men (*P*<0.05) [27].

Spatial hotspot analysis has been utilized in relation with various health policy interventions. For example, Dong et al. (2022) examined the variations of clusters of advanced-stage breast cancer diagnoses in Ohio before and after Medicaid expansion [20]. Dawit et al. (2022) analyzed spatial variations in pre-exposure prophylaxis prescription rates across Ending the HIV Epidemic (EHE) initiative and non-EHE counties in the US [28]. Feng et al. (2023) investigated the factors influencing the adoption of tobacco policy clusters across 351 municipalities in Massachusetts [29]. However, to our knowledge, no study has been conducted to examine the intersection of Medicaid expansion, minimum wage policies, and the hot spots or cold spots of AD dementia prevalence across the US. Our research is motivated by the hypothesis that state-level socioeconomic policies, such as Medicaid expansion and minimum wage, are significantly associated with the distribution of AD dementia. In this study, we identify hot spots and cold spots of AD dementia prevalence across the US and overlay them with state-level Medicaid expansion and minimum wage policy status. Our main objectives are: 1) to assess the association between the counties identified as hot spots and cold spots and the state’s Medicaid expansion status, 2) to evaluate the association between the counties identified as hot spots and cold spots and the state’s minimum wage policy, and 3) to determine whether there are significant differences in the counts of hotspots and cold spots across SVI quartiles.

## 2. Methods

### 2.1. Data

The AD dementia prevalence data was obtained from the Dhana et al. (2023) article, which provides first-ever county-level estimates across the US for adults aged 65 and older [30]. The estimated prevalence was adjusted for age, sex, race/ethnicity, and years of formal schooling [30]. County-level overall SVI data was sourced from the State Cancer Profiles, which draws upon data provided by the Centers for Disease Control and Prevention (CDC). This index measures the resilience of communities when confronted by external stresses on human health [31]. SVI values, a composite score derived from 16 unique census variables, ranged from 0 to 1, with lower values representing lower vulnerability and higher values indicating higher vulnerability [19]. Information on state Medicaid expansion status under the ACA as of April 2019 was obtained from the Kaiser Family Foundation [5]. Data on state minimum wage laws were retrieved from the US Department of Labor [32] and the Economic Policy Institute [33].

### 2.2. Spatial analysis

We assessed the global spatial pattern of AD dementia prevalence across the US using the Getis-Ord General G statistic under the null hypothesis of complete spatial randomness [34]. A higher-than-expected General G with a positive Z-score indicates a clustering of high values. In contrast, a lower-than-expected General G with a negative Z-score indicates an overall tendency to cluster low values [34]. The General G statistic is defined as:

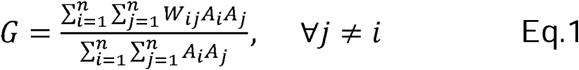

Where *A_i_* and *A_j_* represent AD dementia prevalence rates for counties *i* and *j* respectively; *W_ij_* is the spatial weight between counties *i* and *j* based on the first-order Queen’s contiguity weight matrix (i.e., neighbors if they share a border or vertex); *n* is the total number of counties; and, ∀*j* ≠ *i* ensures the statistic does not consider self-pairs.

The Getis-Ord General G is a global measure of spatial autocorrelation and cannot identify the location of hot or cold spots [35]. Thus, we employed Getis-Ord Gi* statistics to identify local clustering of high (hot spots) or low (cold spots) values of AD dementia prevalence across the US [34]. Using the same notations as Eq.1, the Getis-Ord Gi* statistic for each county is calculated as:

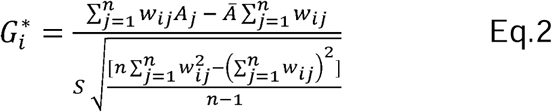

Where *A̅* and *S* represent the mean and standard deviation of AD dementia prevalence across the US, respectively. A significant positive Gi* value above the expected indicates a hot spot, while a negative significant value below the expected indicates a cold spot. We conducted the Getis-Ord analyses using ArcGIS Pro 3.3 (ESRI, Redlands, CA, US) and mapped the outputs with the same software.

### 2.3. Univariate analysis

Based on the Getis-Ord Gi* statistics results, we classified US counties into hot spots, cold spots, and non-significant areas of AD dementia prevalence. The SVI scores were also categorized into quartiles from least vulnerable (Q1) to most vulnerable (Q4). We counted and plotted the number of hot and cold spots that fall into each SVI quartile.

Subsequently, we conducted a Chi-square test to assess whether significant differences exist in the counts of hot and cold spots across SVI quartiles.

Since the Medicaid adoption expansion is decided at the state level [36], we initially calculated the proportion of counties identified as hot and cold spots per state.

Subsequently, we used boxplots to visually compare these proportions categorized by the state’s Medicaid adoption expansion status (adopted vs. not adopted) across the US. We repeated this process for the minimum wage policy and compared these proportions, which were classified by whether the state had a higher minimum wage than the federal level. Mann-Whitney U and Levene’s tests were employed to statistically compare the mean differences and assess the homogeneity of variance between the groups.

### 2.4. Multivariate analysis

We conducted logistic regression analysis on the proportion of hot spot or cold spot counties in each state as the dependent variable. The independent variables were two categorical variables: 1) Medicaid expansion status (adopted vs non adopted) and 2) Minimum wage status (above federal level or not). All statistical analyses were performed in RStudio 2023.06.0 (R Foundation for Statistical Computing, Vienna, Austria).

## 3. Results

The results of the Getis-Ord General G test suggest that the spatial distribution of AD dementia prevalence in the US is more clustered than expected under random spatial processes. The observed General G (0.0322) exceeded the expected General G (0.0319), with a significant *Z*-score (15.77) and a small p-value (*P*<0.001), indicating a robust spatial autocorrelation and significant clustering of high values. The Getis-Ord Gi* statistics further identified hot spots and cold spots of AD dementia prevalence at 90%, 95%, and 99% confidence levels. The hot spots were primarily located in the Deep South states, with approximately 15.5% of US counties (n=480 out of 3,105) (*P*<0.05). In contrast, the cold spots were mainly found in western and northeastern states, with additional counties in Kentucky, Alaska, northern Michigan, and northern Wisconsin. Approximately 14.8% of US counties (n=460 out of 3,105) were identified as cold spots (*P*<0.05). However, most counties in central and northern states were neither hot spots nor cold spots. Figure 1 illustrates the spatial distribution of hot spots, cold spots, and non-significant areas of AD dementia prevalence in the US.

**Fig 1.**
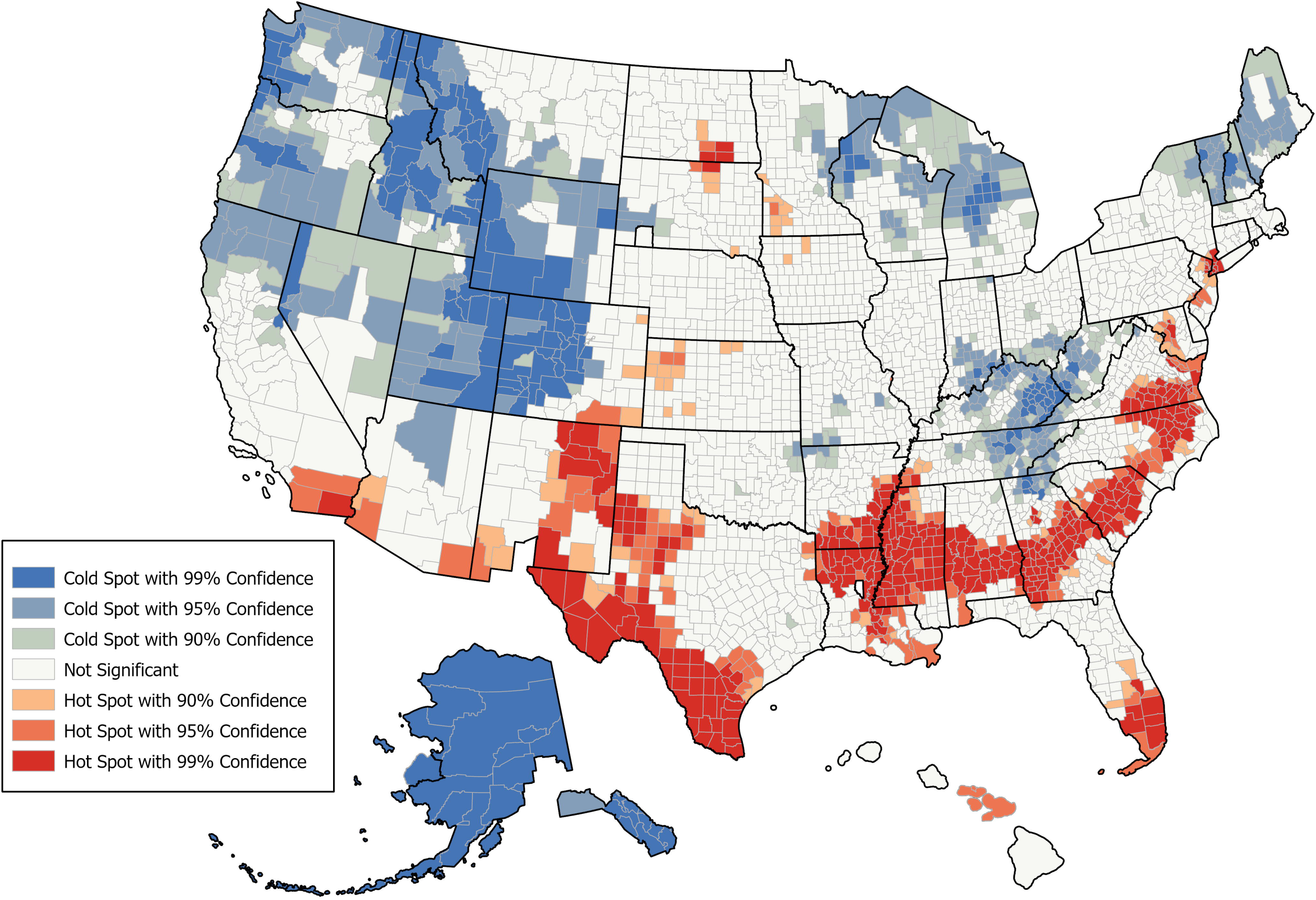
Geospatial distribution of hot spots, cold spots, and non-significant areas of AD dementia prevalence identified by Getis-Ord Gi* across the US at various confidence levels.

A comparison of SVI quartiles with the location of hot spot counties revealed that the most vulnerable SVI quartile (Q4) had the highest number of hot spots (n=311, 64.8% of all hot spots), followed by Q3 (n=101, 21.0% of all hot spots), and Q2 (n=46, 9.6% of all hot spots). The least vulnerable SVI quartile (Q1) had the fewest hot spots (n=22, 4.6%). Conversely, the comparison of SVI quartiles with the location of cold spots indicated that the most vulnerable SVI quartile (Q4) had the fewest cold spots (n=54, 11.7% of all cold spots). The highest number of cold spots were in Q2 (n=150, 32.6%), followed by Q3 (n=138, 30.0%) and Q1 (n=118, 25.7%). Moreover, the difference was statistically significant (,²=307.41, df = 3, *P*<0.01). Figure 2 illustrates the frequency of hot spots and cold spots of AD dementia prevalence in the US, classified by SVI quartiles.

**Fig 2.**
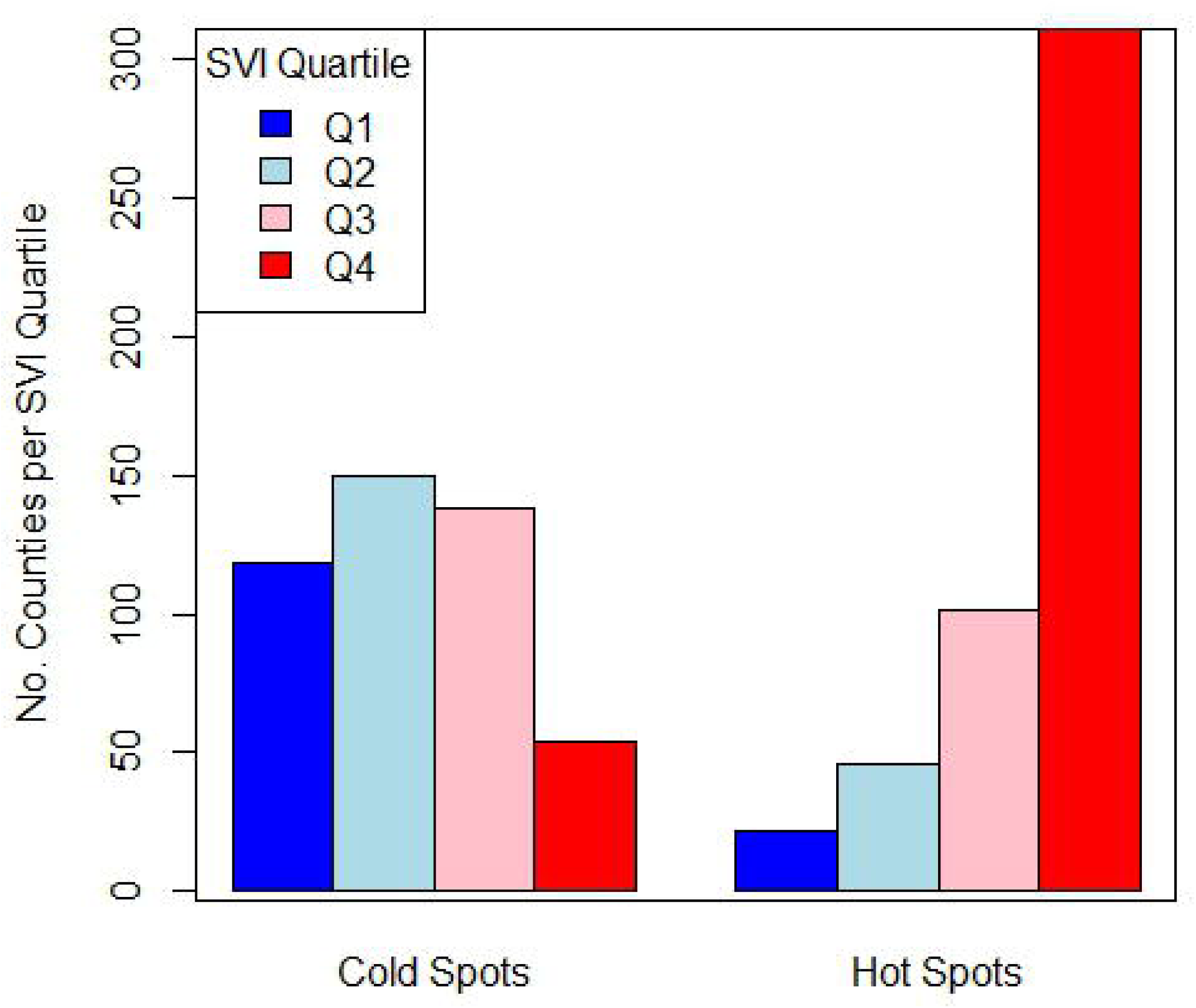
The frequency of hot spot and cold spot counties of AD dementia prevalence in the US, classified by SVI quartiles.

The comparison of hot spot proportion per state with Medicaid expansion status showed that the states that adopted the Medicaid expansion had a lower proportion of hot spots than non-adopting states. Mann-Whitney U test showed that this difference was significant (*P*<0.05). Additionally, the adopting states exhibited smaller variability (more consistent pattern) in the proportion of hot spots than non-adopting (Figure 3A).

**Fig 3.**
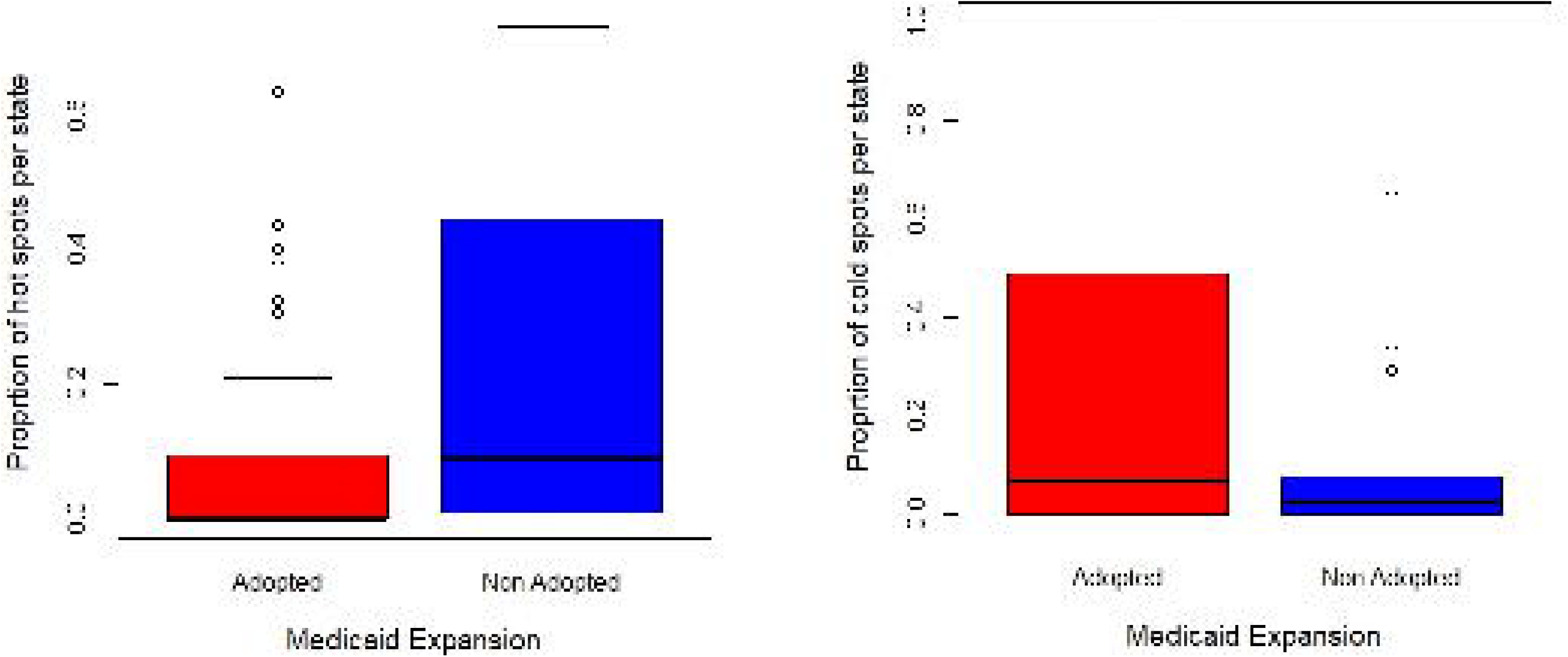
Boxplots showing the proportion of (A) hot spot and (B) cold spot counties per state, categorized by Medicaid expansion status.

Levene’s test indicated unequal variances between these two groups (*F*=5.6, *P*<0.05). In contrast, when comparing the proportion of cold spots per state, adopting states had a higher proportion of cold spots than those that did not; however, the difference was not statistically significant (*P*>0.05). The adopting states demonstrated greater variability in the proportion of cold spots (Figure 3B), and the difference was significant at a 90% confidence level (*F*=3.0, *P*<0.1). Table 1 summarizes the statistics on the proportion of hot and cold spots per state according to Medicaid expansion status.

**Table 1.** Summary statistics of the proportion of hot and cold spot counties per state classified by the Medicaid expansion status.

|  | Medicaid<br>Expansion | Mean | Median | Standard<br>Deviation | Interquartile<br>Range |
| --- | --- | --- | --- | --- | --- |
| **Proportion**<br>**of Hot spots** | Adopted | 0.09 | 0.00 | 0.16 | 0.08 |
|  | Non-adopted | 0.23 | 0.09 | 0.26 | 0.41 |
| **Proportion**<br>**of Cold**<br>**spots** | Adopted | 0.24 | 0.07 | 0.30 | 0.48 |
|  | Non-adopted | 0.11 | 0.02 | 0.19 | 0.06 |

The comparison of hot spots proportion per state with the minimum wage policy suggests that the states with higher minimum wage than the federal level had a lower proportion of hot spots than those at or below minimum wage than the federal level. However, the Mann-Whitney U test showed that this difference was insignificant (*P*>0.05). Additionally, the higher minimum wage states exhibited smaller variability in the proportion of hot spots (Figure 4A). Levene’s test indicated unequal variances between these two groups at a 90% confidence level (*F*=3.2, *P*<0.1). In contrast, when comparing the proportion of cold spots per state, higher minimum wage states showed almost a similar proportion of cold spots than at or below the minimum wage states. The mean and variance differences were not significantly different (*P*>0.05). Table 2 summarizes the statistics on the proportion of hot and cold spots per state according to the minimum wage policy.

**Fig 4.**
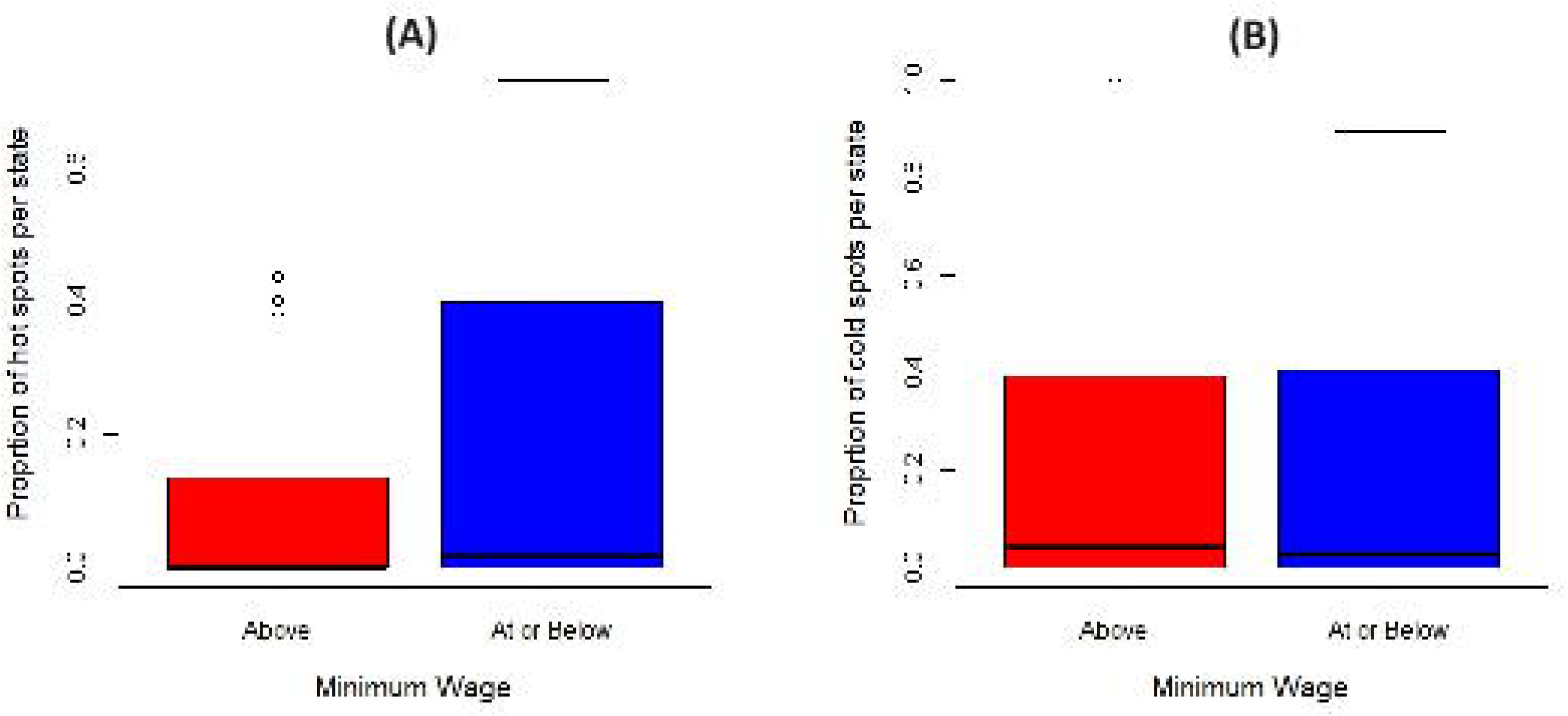
Boxplots showing the proportion of (A) hot spot and (B) cold spot counties per state, categorized by the minimum wage policy.

**Table 2.**
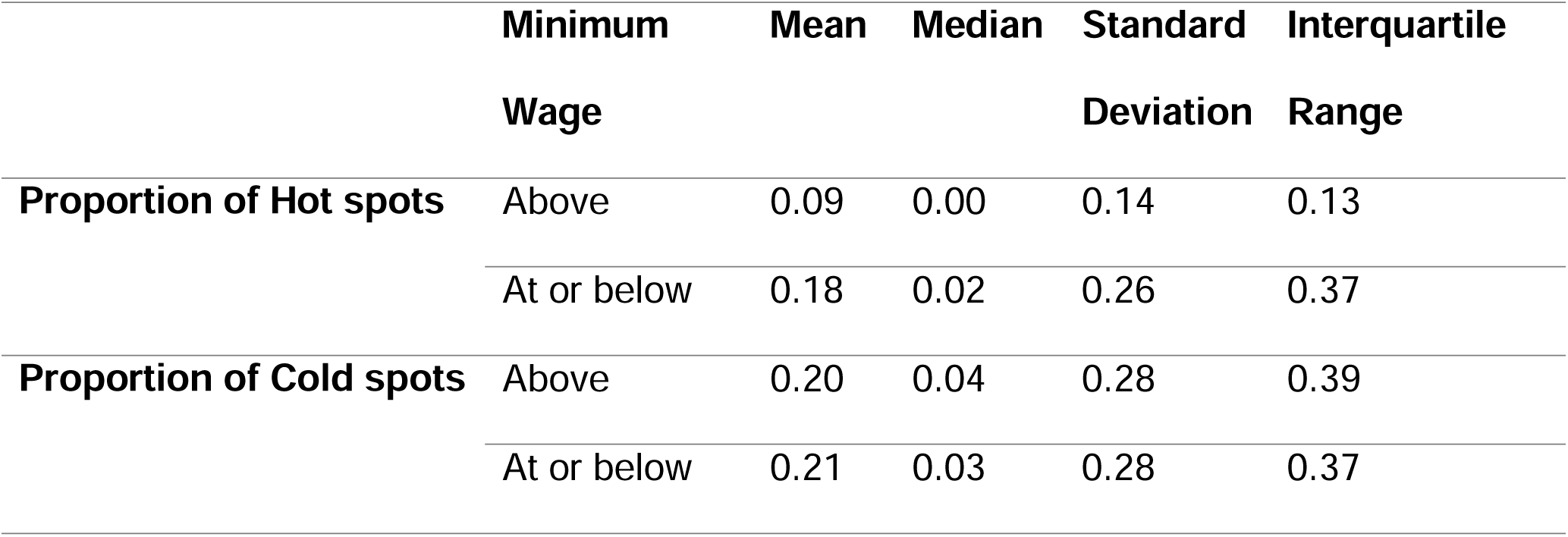
Summary statistics of the proportion of hot and cold spots per state by minimum wage policy.

Logistic regression was performed to examine the association between the count of hot spots vs. non-hot spots counties per state, considering Medicaid adoption status and minimum wage policy as predictors. The non-adopting states had significantly higher odds of having hot spots compared to adopting states (OR=2.58, 95% CI:2.04–3.26, *P*<0.001). Additionally, the lower minimum wage states showed significantly higher odds of having hot spots of AD dementia prevalence than higher minimum wage states (OR=1.94, 95% CI:1.51–2.49, *P*<0.01). Similarly, a logistic regression model was fitted to examine the relationship between the count of cold spots vs. non-cold spots counties per state, considering Medicaid adoption status and minimum wage policy as predictors. The non-adopting states had significantly lower odds of having cold spots than the adopting states (OR=0.24, 95% CI: 0.19–0.32, *P*<0.01). However, the lower minimum wage states exhibited significantly higher odds of cold spots than higher minimum wage states (OR=1.45, 95% CI:1.16–1.81, *P*<0.05).

## 4. Discussion

To support the National Plan to Address Alzheimer’s Disease goals [37], our study aimed to provide geospatial insights into the nationwide distribution of AD dementia prevalence in relation to Medicaid expansion, minimum wage policies, and overall SVI scores. Based on our findings, a higher proportion of hotspots were found in states that did not adopt Medicaid expansion, had minimum wage policies at or below the federal level, were located in the Deep South, and were in the most vulnerable SVI quartiles. Conversely, cold spots were predominantly located primarily in states that did adopt Medicaid expansion, had minimum wage policies above the federal level, were located in Western and Northeastern regions, and fell into the least vulnerable SVI quartiles. These findings provide geospatial evidence into the relationship between governmental policies and social determinants of health with the prevalence of AD dementia in the US. Previous geospatial studies of AD across the US have identified different hotspot locations. For instance, Xu et al. (2018) used individual death certificate data and space-time scan statistics to identify hotspots of AD and all-cause dementia mortality from 2000 to 2010, finding that hotspots were mainly located in the Carolinas, the Ohio River Valley, and the Pacific Northwest [38]. In another study, Amin et al. (2018) used the CDC Wonder Multiple Cause of Death database and spatial scan statistics, identifying age-adjusted AD mortality hotspots from 2008 to 2012 with hotspots primarily in Washington, Iowa, and the Dakotas [39]. More recently, Li et al. (2024) used inpatient claims data for Medicare fee-for-service beneficiaries and space-time scan statistics to identify the hotspots of comorbid cancer and dementia from 2013 to 2018, locating them in the Middle and South Atlantic regions and the Midwestern US [25]. The variations in hotspot locations across these studies may be attributed to differences in data sources, the type of data (prevalence vs mortality), the method used, and the study periods.

However, the data sources utilized in our study utilized the first-ever county-level estimates of AD dementia prevalence in the US. The data providers emphasized the distinct advantages of their estimates over conventional methods, which often rely on medical reports, insurance claims, and national surveys. These traditional methods frequently underestimate prevalence due to data discrepancies and systemic diagnostic biases, particularly affecting racial and ethnic minority groups [30].

When considering governmental policies that address care coverage, the Medicaid expansion program is critically important for people living with AD dementia. As AD dementia progresses, individuals increasingly rely on paid care assistance for long-term support services, which are not covered by Medicare, resulting in high out-of-pocket expenses that low-income individuals cannot afford [40]. However, Medicaid does cover long-term support services [41], making it a crucial resource for those in need. People living with AD dementia frequently qualify for Medicaid based on income and disability, and some states have extended eligibility through pathways targeting people with disabilities or long-term support service needs. Despite these provisions, not all low-income people living with AD dementia qualify for Medicaid, leaving many struggling to secure the necessary assistance. This gap in coverage makes access to adequate long-term support services more essential, as these services help to prevent adverse outcomes related to safety (falls, wandering, harm), nutrition (forgetting to eat, aspiration), self-care (poor hygiene leading to secondary infections), and pain (decreased mobility, contractures, pressure sores) [42]. Conversely, areas with better access to care often benefit from enhanced preventative and early intervention services that can manage or delay the progression of symptoms, thereby reducing the number of AD diagnoses. Comprehensive care settings might result in diagnosing other conditions with similar symptoms, potentially influencing the diagnostic rates.

The relationship between higher minimum wages and increased AD prevalence may be linked to underlying socioeconomic factors rather than the wage level itself. In states that have not adopted the Medicaid expansion, a significant proportion of low-income individuals living with AD dementia likely do not qualify for Medicaid [43]. Our findings suggest that AD dementia prevalence is prevalent in areas that have limited access to Medicaid, lower minimum wages, and higher social vulnerability. This suggests that a significant percentage of low-income individuals with AD dementia in these areas may not have access to the long-term support services they need. States may have programs outside of Medicaid to support the long-term care needs of people living with AD dementia. Future studies should explore these alternative programs and assess the extent of service access issues in these states.

Unlike the recent Medicaid expansion program, the government has established a minimum wage as a long-standing policy. Prior research has established that prolonged exposure to low minimum wage [44] and lower household income [45, 46] is associated with an increased risk of AD dementia; our geospatial analysis supports this relationship. However, the causal relationship between these factors is still unclear. The relationship is likely mediated by many factors. Low-wage jobs are associated with factors known to increase the risk for AD dementia, including toxic exposures, risk of injuries and trauma [47], obesity [48], and factors associated with low cognitive reserve (e.g., depression, low cognitive stimulation, limited diversity in lived experiences, and high stress) [49, 50]. It is also noteworthy that minimum wage laws often correlate with the cost of living; higher living costs typically prompt states or localities to set higher minimum wages. Further causal analyses are needed to explore the direct impact of these factors on the geospatial pattern observed for AD dementia prevalence.

Several limitations should be noted. The cross-sectional nature of this study and the lack of longitudinal data preclude establishing causality. Further, there may be a lag between policy adoption and implementation in certain states, which can bias the attribution of observed patterns to those policies. Moreover, other influential contextual factors, such as healthcare access and the cultural and political leanings of the states, were not explored, potentially confounding or modifying the observed relationships.

Future studies must conduct causality analysis to uncover the underlying mechanisms through which the policies might exert their impacts. Additionally, different specifications of hot/cold spot detection techniques may yield varying results, highlighting the need for conducting further robust sensitivity analyses.

## 5. Conclusions

In summary, this nationwide study offers foundational geospatial insights into the distribution of AD dementia prevalence, highlighting significantly impacted areas associated with high SVI scores, minimal minimum wage policies, and states not adopting Medicaid expansion. The findings suggest there may be barriers to care access for low-income individuals with AD dementia in states that have not adopted the Medicaid expansion that need to be examined. Additionally, future studies should examine the causal relationship between low minimum wage policies and the risk of AD dementia, taking into consideration varying lengths of exposure. Together, these insights provide a baseline for more nuanced research into the impact of economic policies on geospatial patterns for people living with AD.

## Data Availability

The original contributions presented in the study are included in the article, further inquiries can be directed to the corresponding author.

